# Association between Participation in Government Subsidy Program for Domestic Travel and Symptoms Indicative of COVID-19 Infection

**DOI:** 10.1101/2020.12.03.20243352

**Authors:** Atsushi Miyawaki, Takahiro Tabuchi, Yasutake Tomata, Yusuke Tsugawa

## Abstract

**Importance:** As countermeasures against the economic downturn caused by the coronavirus 2019 (COVID-19) pandemic, many countries have introduced or considering financial incentives for people to engage in economic activities such as travel and use restaurants. Japan has implemented a large-scale, nationwide government-funded program that subsidizes up to 50% of all travel expenses since July 2020 with the aim of reviving the travel industry. However, it remains unknown as to how such provision of government subsidies for travel impacted the COVID-19 pandemic.

**Objective:** To investigate the association between participation in government subsidies for domestic travel in Japan and the incidence of COVID-19 infections.

**Design, Setting, and Participants:** Using the data from a large internet survey conducted between August 25 and September 30, 2020, in Japan, we examined whether individuals who used subsidies experienced a higher likelihood of symptoms indicative of the COVID-19 infection.

**Exposure:** Participation in the government subsidy program for domestic travel.

**Main Outcomes and Measures:** Five symptoms indicative of the COVID-19 infection (high fever, sore throat, cough, headache, and smell and taste disorder) within the past one month of the survey.

**Results:** Of the 25,482 respondents (50.3% [12,809] women; mean [SD] age, 48.4 [17.4] years), 3,289 (12.9%) participated in the subsidy program at the time of survey. After adjusting for potential confounders, we found that participants in the subsidy program exhibited higher incidence of high fever (adjusted rate, 4.8% for participants vs. 3.7% for non-participants; adjusted odds ratio [aOR], 1.90; 95%CI, 1.42-2.54; p<0.001), sore throat (19.8% vs. 11.3%; aOR, 2.09; 95%CI, 1.37-3.20; p=0.002), cough (19.1% vs. 11.2%; aOR 1.96; 95%CI, 1.27-3.02; p=0.007), headache (29.1% vs. 25.5%; aOR, 1.24; 95%CI, 1.07-1.43; p=0.007), and smell and taste disorder (2.6% vs. 1.8%; aOR 1.98; 95%CI; 1.15-3.40; p=0.01) compared with non-participants.

**Conclusion and Relevance:** The participants of government subsidies for domestic travel experienced a higher incidence of symptoms indicative of the COVID-19 infection.

## INTRODUCTION

As of the end of November 2020, 62 million people have been infected by the coronavirus disease 2019 (COVID-19), and 1.4 million have died of this infection.^1^ In order to tackle this unprecedented pandemic, many countries have implemented public health measures — also known as non-pharmaceutical interventions (NPIs) — to control the spread of the virus, including lockdown, movement restrictions, quarantine, and border control.^2^ Given that the number of infections and deaths due to COVID-19 has been resurging during this winter, these NPIs are likely to be implemented intermittently,^3^ until effective vaccines would be developed and become widely available. While these NPIs have been shown to be effective in reducing the spread of COVID-19 infections,^2,4^ they have a substantial negative impact on the economy.^5^ To countermeasure the economic downturn due to the NPIs, many countries have introduced or actively considering financial incentives such as government subsidies to engage in economic activities such as to use restaurants or travel domestically.^6–9^

Evidence is limited as to whether the government interventions to financially incentivize economic activities such as using restaurants or traveling impact the COVID-19 infection. For example, the United Kingdom implemented the “*Eat out to Help out*” campaign, in which the government subsidized up to 50% of the expenses of food and non-alcoholic drinks for immediate consumption at restaurants using a budget of around £500 million throughout August.^9^ A recent study using ecological data on COVID-19 infections by region suggested that regions that implemented this campaign experienced 8-17 percentage points higher number of COVID-19 clusters.^10^ However, ecological association does not imply that the same association could be observed at the individual level (known as “ecological fallacy”), and therefore, it remains unknown as to whether this policy actually led to an increased number of individuals infected by the COVID-19. Indeed, to our knowledge, no study to date has evaluated the impact of such economic policy on the risk of the COVID-19 infection using individual-level data. Moreover, it remains unknown as to how similar policies implemented in other countries that incentivize economic activities (e.g., eating out, travel) affected the COVID-19 pandemic.

Japan has implemented large-scale, nationwide government subsidies for domestic travel (called the “*Go-To*” Domestic Travel Campaign)^8^ since July 2020 with the aim of reviving the travel industry, which has been hit hard by a substantial decrease in the number of foreign tourists visiting Japan. This program incentivizes people to travel domestically by subsidizing up to 50% of all travel expenses for those who travel, including transport and accommodation expenses. As of the end of October 2020, more than 200 billion Japanese yen (JPY) (approximately 2 billion US dollars, using an exchange rate of 100 JPY per US dollar) have been used to subsidize a total of 40 million people who traveled domestically.^11^ However, as the number of the COVID-19 infected cases has resurged, the Japanese government is facing fierce criticisms speculating that increased mobility and human interactions due to this program may be causing the increase in the number of COVID-19 infections.^8^ Yet, empirical evidence is lacking as to whether the introduction of this program is associated with an increased risk of the COVID-19 infection. Japan’s experience from this social experiment provides a unique opportunity to understand the impact of government subsidies for travel on the spread of COVID-19 infections.

In this context, using the data from a large internet survey conducted between August 25 and September 30, 2020, in Japan, we examined whether individuals who used subsidies experienced a higher incidence of symptoms indicative of the COVID-19 infection (COVID-19-like symptoms).

## METHODS

### Study Design, Setting, and Data Sources

We analyzed the data from the *Japan “COVID-19 and Society” Internet Survey (JACSIS)* study, a cross-sectional, web-based, self-reported questionnaire survey administered by a large internet research agency (Rakuten Insight, Inc., which had approximately 2.2 million qualified panelists in 2019).^12^ This internet research agency has been used in previous studies.^13,14^ The questionnaires were distributed to 224,389 panelists selected by each sex, age, and prefecture category using simple random sampling (it covered all 47 prefectures, the first-tier administrative district in Japan). The panelists who consented to participate in the survey accessed the designated website and responded to the questionnaires, and had the option not to respond or discontinue at any point of the survey. The questionnaires were distributed starting on August 25, 2020, and was completed on September 30, 2020, when the target numbers of respondents for each sex, age, and prefecture category were met (the target numbers for each sex, age, and prefecture category had been determined in advance according to the population distribution in 2019; 28000 respondents; response rate, 12.5%). We excluded 2,518 individuals showing unnatural or inconsistent responses using the algorithm we developed. The final sample size was 25,482 respondents (91.0% of the total survey respondents).

### Exposure Variables

The primary exposure variable was the participation in the subsidy program for domestic travel, which has been effective since July 22, 2020.

### Outcome Variables

Our outcome variable was the incidence of five self-reported COVID-19-like symptoms (high fever, sore throat, cough, headache, and smell and taste disorder) during the one month prior to the time of the survey.^15^ Self-reported COVID-19-like symptoms have been reported as a useful measure to monitor the spread of the COVID-19 infections.^16,17^

### Adjustment Variables

We adjusted for the respondents’ demographics,^18^ socio-economic status (SES),^19^ health-related characteristics,^18^ and prefecture fixed effects (effectively comparing individuals living in the same prefecture). The demographics included age (categorized as 15-19, 20-29, …, 70-79) and sex. The SES included academic attainment (graduated from college or higher institutions vs. high school or lower institutions), income level (categorized using the tertiles of household equivalent income [“low” = less than 2.5 million JPY, “medium” = 2.5 to 4.3 million JPY, and “high” = more than 4.3 million JPY], and an indicator for those who refused to respond to this question), household size (number of household members: 1, 2, 3, 4 and 5+), employment status (employer, self-employed, employee, and unemployed), and marital status (married, never married, widowed, and separated). The household equivalized income was calculated as the gross (pre-tax) income in 2019, divided by the square root of the number of household members. Health-related characteristics included smoking status (never, ever, and current smokers), walking disability (whether the person is experiencing difficulties in walking), and eight comorbidities (overweight [body mass index ≥ 25 kg/m^2^], hypertension, diabetes, asthma, coronary disease, stroke, chronic obstructive pulmonary disease, and cancer). Body mass index was calculated by dividing self-reported body weight by self-reported body height squared (m^2^).

### Statistical Analysis

First, we compared the demographics, SES, and health-related characteristics of the participants in the subsidy program for domestic travel vs. non-participants. To account for the possibility that those who participated and responded to the internet-based survey may differ from the general population (e.g., a younger population may be more likely to participate and respond to the internet-based survey), we used the weighted regression models (inverse probability weighting [IPW]).^20^ The weights (propensity scores) were calculated by fitting a logistic regression model using demographics, SES, and health-related characteristics to adjust for the difference in respondents between the current internet survey and a widely-used nationwide representative survey (i.e., the 2016 Comprehensive Survey of Living Conditions^21,22^) (see **Supplementary Method 1** for details).

Second, we examined the association between participation in the subsidy program for domestic travel and the incidence rates of the COVID-19-like symptoms. For each outcome, we adjusted for the respondents’ sociodemographic characteristics, health-related characteristics, and prefecture fixed effects. We used weighted multivariable logistic regression models, with standard errors clustered at the prefecture-level, to account for the potential correlation of the respondents within the same prefecture. To calculate risk-adjusted incidence rates of COVID-19-like symptoms, we used marginal standardization (also known as predictive margins or margins of response).^23^ For each respondent, we calculated predicted probabilities of the incidence of the COVID-19-like symptom with participation in the subsidy program fixed at each category and then averaged over the distribution of covariates in our sample.

To adjust for multiple comparisons of having five outcome variables using the Holm method,^24^ which sequentially compares the *i*-th smallest P value (for *i* = 1, …, 5) among the five original P values with progressively less restrictive alpha levels (= .05/(5 − *i* + 1)). To make the interpretation easier, we calculated the adjusted P value by multiplying the unadjusted P values by (5 – *i* + 1) times, and considered the adjusted P value < 0.05 to be statistically significant.^25^ All analyses were conducted using Stata version 15 (College Station, TX; StataCorp LLC.). This study was approved by the Institutional Review Board of the Osaka International Cancer Institute (No. 20084).

### Secondary analysis

We conducted sensitivity analyses. First, the travels to and from Tokyo were ineligible for the subsidy program until September 15, due to a large number of the COVID-19 cases in Tokyo.^8^ To assess whether our findings were sensitive to the inclusion of residents of Tokyo (we did not exclude these individuals in our main analyses as they could still use the subsidy program if their companion lived in other prefectures than Tokyo), we reanalyzed the data after excluding the respondents living in Tokyo prefecture. Second, we repeated the analyses without using IPW to examine how the use of this approach affected our findings. Third, in order to test whether the impact of the subsidy program differs by the characteristics of respondents, we conducted stratified analyses by age (15-64 years and 65-79 years), the presence of comorbidities (no comorbidities vs. having at least one comorbidity), and sex.

## RESULTS

### Characteristics of respondents

Of the 25,482 respondents, 3,289 (12.9%) had participated in the subsidy program for domestic travel at the time of the survey. The participants in the subsidy program were younger; have higher education and higher income; and more likely to be overweight (**Table 1**).

**Table 1.**
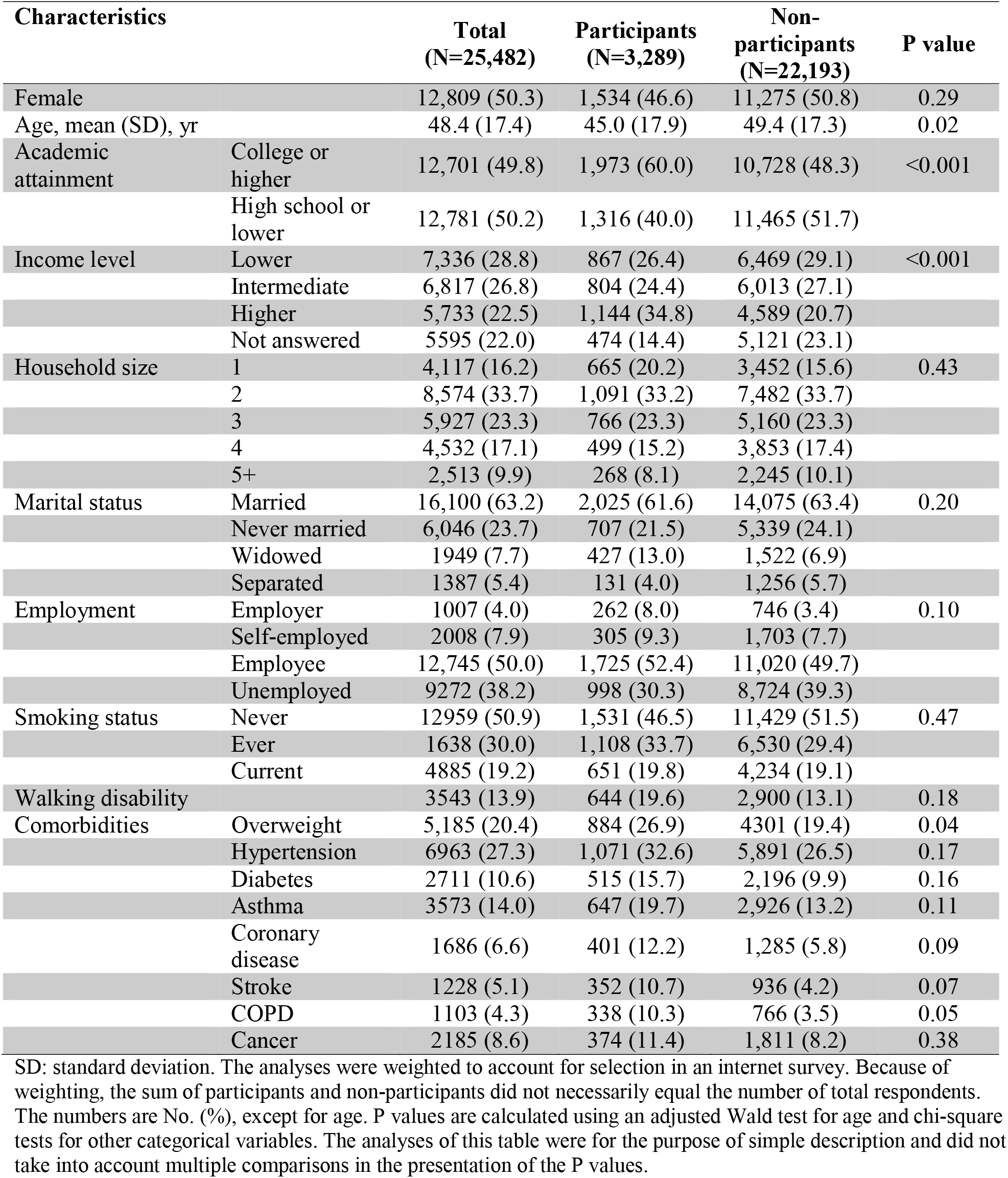
Sociodemographic and Health-Related Characteristics of Respondents by Participation in the Subsidy Program for Domestic Travel.

| Characteristics |  | Total<br>(N=25,482) | Participants<br>(N=3,289) | Non-participants<br>(N=22,193) | P value |
| --- | --- | --- | --- | --- | --- |
| Female |  | 12,809 (50.3) | 1,534 (46.6) | 11,275 (50.8) | 0.29 |
| Age, mean (SD), yr |  | 48.4 (17.4) | 45.0 (17.9) | 49.4 (17.3) | 0.02 |
| Academic attainment | College or higher | 12,701 (49.8) | 1,973 (60.0) | 10,728 (48.3) | <0.001 |
|  | High school or lower | 12,781 (50.2) | 1,316 (40.0) | 11,465 (51.7) |  |
| Income level | Lower | 7,336 (28.8) | 867 (26.4) | 6,469 (29.1) | <0.001 |
|  | Intermediate | 6,817 (26.8) | 804 (24.4) | 6,013 (27.1) |  |
|  | Higher | 5,733 (22.5) | 1,144 (34.8) | 4,589 (20.7) |  |
|  | Not answered | 5595 (22.0) | 474 (14.4) | 5,121 (23.1) |  |
| Household size | 1 | 4,117 (16.2) | 665 (20.2) | 3,452 (15.6) | 0.43 |
|  | 2 | 8,574 (33.7) | 1,091 (33.2) | 7,482 (33.7) |  |
|  | 3 | 5,927 (23.3) | 766 (23.3) | 5,160 (23.3) |  |
|  | 4 | 4,532 (17.1) | 499 (15.2) | 3,853 (17.4) |  |
|  | 5+ | 2,513 (9.9) | 268 (8.1) | 2,245 (10.1) |  |
| Marital status | Married | 16,100 (63.2) | 2,025 (61.6) | 14,075 (63.4) | 0.20 |
|  | Never married | 6,046 (23.7) | 707 (21.5) | 5,339 (24.1) |  |
|  | Widowed | 1949 (7.7) | 427 (13.0) | 1,522 (6.9) |  |
|  | Separated | 1387 (5.4) | 131 (4.0) | 1,256 (5.7) |  |
| Employment | Employer | 1007 (4.0) | 262 (8.0) | 746 (3.4) | 0.10 |
|  | Self-employed | 2008 (7.9) | 305 (9.3) | 1,703 (7.7) |  |
|  | Employee | 12,745 (50.0) | 1,725 (52.4) | 11,020 (49.7) |  |
|  | Unemployed | 9272 (38.2) | 998 (30.3) | 8,724 (39.3) |  |
| Smoking status | Never | 12959 (50.9) | 1,531 (46.5) | 11,429 (51.5) | 0.47 |
|  | Ever | 1638 (30.0) | 1,108 (33.7) | 6,530 (29.4) |  |
|  | Current | 4885 (19.2) | 651 (19.8) | 4,234 (19.1) |  |
| Walking disability |  | 3543 (13.9) | 644 (19.6) | 2,900 (13.1) | 0.18 |
| Comorbidities | Overweight | 5,185 (20.4) | 884 (26.9) | 4301 (19.4) | 0.04 |
|  | Hypertension | 6963 (27.3) | 1,071 (32.6) | 5,891 (26.5) | 0.17 |
|  | Diabetes | 2711 (10.6) | 515 (15.7) | 2,196 (9.9) | 0.16 |
|  | Asthma | 3573 (14.0) | 647 (19.7) | 2,926 (13.2) | 0.11 |
|  | Coronary disease | 1686 (6.6) | 401 (12.2) | 1,285 (5.8) | 0.09 |
|  | Stroke | 1228 (5.1) | 352 (10.7) | 936 (4.2) | 0.07 |
|  | COPD | 1103 (4.3) | 338 (10.3) | 766 (3.5) | 0.05 |
|  | Cancer | 2185 (8.6) | 374 (11.4) | 1,811 (8.2) | 0.38 |

SD: standard deviation. The analyses were weighted to account for selection in an internet survey. Because of weighting, the sum of participants and non-participants did not necessarily equal the number of total respondents. The numbers are No. (%), except for age. P values are calculated using an adjusted Wald test for age and chi-square tests for other categorical variables. The analyses of this table were for the purpose of simple description and did not take into account multiple comparisons in the presentation of the P values.

### Participation in the subsidy program for domestic travel and COVID-19-like symptoms

After adjusting for demographics, SES, health-related characteristics and indicators of prefectures (**Table 2**), we found that the adjusted incidence rates of COVID-19-like symptoms were higher for the subsidy program participants compared with the non-participants for high fever (adjusted rate, 4.8% for participants vs. 3.7% for non-participants; adjusted odds ratio [aOR], 1.90; 95%CI, 1.42-2.56; p<0.001), sore throat (19.8% vs. 11.3%; aOR, 2.09; 95%CI, 1.37-3.20; p=0.002), cough (19.1% vs. 11.2%; aOR 1.96; 95%CI, 1.27-3.02; p=0.007), headache (29.1% vs. 25.5%; aOR, 1.24; 95%CI, 1.07-1.43; p=0.007), and smell and taste disorder (2.6% vs. 1.8%; aOR 1.98; 95%CI; 1.15-3.40; p=0.01).

**Table 2.** Association between Participation in the Subsidy Program for Domestic Travel and Incidence of COVID-19-Like Symptoms.

| Subsidy Program Participation | Weighted sample, No. | Weighted incidence, n (%) | Adjusted rate, % (95% CI) | Adjusted OR (95% CI) | Adjusted P value |
| --- | --- | --- | --- | --- | --- |
| High Fever |  |  |  |  |  |
| Participants | 3,289 | 327 (9.9) | 4.8 (4.2, 5.3) | 1.90 (1.42, 2.54) | <0.001 |
| Non-participants | 22,193 | 633 (2.9) | 3.7 (3.6, 3.8) | Reference |  |
| Sore Throat |  |  |  |  |  |
| Participants | 3,289 | 790 (24.0) | 19.8 (15.1, 24.6) | 2.09 (1.37, 3.20) | 0.002 |
| Non-participants | 22,193 | 2406 (10.8) | 11.3 (10.5, 12.1) | Reference |  |
| Cough |  |  |  |  |  |
| Participants | 3,289 | 728 (22.1) | 19.1 (14.3, 24.0) | 1.96 (1.27, 3.02) | 0.007 |
| Non-participants | 22,193 | 2417 (10.9) | 11.2 (10.5, 12.0) | Reference |  |
| Headache |  |  |  |  |  |
| Participants | 3,289 | 1,009 (30.7) | 29.1 (26.9, 31.3) | 1.24 (1.07, 1.43) | 0.007 |
| Non-participants | 22,193 | 5,612 (25.3) | 25.5 (25.2, 25.8) | Reference |  |
| Smell and Taste Disorder |  |  |  |  |  |
| Participants | 3,289 | 167 (5.1) | 2.6 (2.0, 3.1) | 1.98 (1.15, 3.40) | 0.01 |
| Non-participants | 22,193 | 287 (1.3) | 1.8 (1.6, 1.9) | Reference |  |

OR: odds ratio. CI: confidence interval. We examined the association of the participation in the government subsidies for domestic travel in the past 1-2 months with the incidence of the five COVID-19-like symptoms in the past one month. For each outcome, we constructed a weighted multivariable logistic regression model with standard errors clustered at the prefecture-level. We adjusted for the respondents' sociodemographic characteristics, health-related characteristics, and prefecture indicator variables. We weighted the regression models using IPW to account for "being a respondent in an internet survey." Adjusted rates were calculated using marginal standardization. Adjusted P values using the Holm method for multiple testing were shown (the adjusted p value < 0.05 was considered to be statistically significant).

### Secondary analysis

Our findings were largely unaffected by excluding the respondents living in Tokyo (**Supplementary Table 1**) or using unweighted regression models (**Supplementary Table 2**). The result of the stratified analyses by age showed a higher incidence rate of COVID-19-like symptoms were more salient among young respondents (**Supplementary Table 3**). For example, among respondents aged 15-64 years, the adjusted incidence rate of smell and taste disorder was higher for the subsidy program participants compared with the younger non-participants (adjusted rate, 3.4% vs. 2.4%; aOR, 2.00; 95%CI, 1.15-3.48; p=0.01), whereas the incidence rates did not differ between participants and non-participants among those aged 65-79 years (0.3% vs. 0.6%; aOR, 0.52; 95%CI, 0.20-1.38; p=0.19) (p for interaction = 0.02). We found no systemic difference in the patterns regarding the association between the subsidy program participation and COVID-19-like symptoms for the stratified analyses by the presence of comorbidity and sex (**Supplementary Tables 4-5**).

## DISCUSSION

Using the data from a large cross-sectional internet survey that included more than 25,000 adults in Japan, we found that individuals who participated in the government’s subsidy program for domestic travel experienced a higher incidence of COVID-19-like symptoms compared with those who did not participate. This association was also observed for the incidence of smell and taste disorder, which is a highly specific symptom of the COVID-19 infection.^15,26^ This increased incidence of the COVID-19-like-symptoms was clustered among individuals aged <65 years, but not for those aged ≥ 65 years, suggesting that, if any, the non-elderly generation may be contributing to the spread of COVID-19 infection associated with this program. Given that the Japanese government is currently considering to halt this subsidy program with concerns for increased risks of COVID-19 infections, and other countries are also actively considering similar policies to stimulate the economy, our findings should be informative for policymakers to design policies that could increase economic activities without exacerbating the COVID-19 pandemic. There are several mechanisms through which participation in this subsidy program was associated with a higher incidence of COVID-19-like symptoms. First, traveling itself may have led to a higher risk of incidence of COVID-19 due to increased contact with people in dining and sightseeing at the destination (causal effect). This explanation is supported by a recent genome epidemiological study of SARS-CoV-2 in Japan that found the possibility that the COVID-19 clusters in Tokyo metropolitan areas might have spread throughout Japan after the lifting of movement restrictions.^27^ Second, the subsidy program participants might have been more likely to take behaviors that place them at greater risk of contracting the COVID-19 than the non-participants (selection effect). However, even if the findings were to be explained by this selection effect, our findings nevertheless indicate that the subsidy program may be incentivizing those who had higher risks of COVID-19 transmission to travel, leading to larger cases of infections. A better policy may be that incentivize individuals with a lower risk of the COVID-19 infection to travel, while those with a high risk to stay at home.

Our study has limitations. First, as with any observational study, we could not fully account for unmeasured confounders, and our study was unable to identify the exact mechanisms of the association between the subsidy program participation and increased incidence rates of COVID-19-like symptoms. Second, given the cross-sectional design of our study, we could not identify the temporal relationship between the subsidy program and the incidence of the COVID-19-like symptoms. Instead of the government subsidy causing the infection of the COVID-19, it was also possible that individuals who had experienced COVID-19-like symptoms were more likely to utilize the program and travel domestically. However, this explanation may be unlikely given that travel agents and hotels have been introducing strict protocols to ensure that nobody with the COVID-19-like symptoms to use their services, and individuals who spread the virus are likely to face criticism and stigma in Japan incentivizing people with suspected symptoms to stay at home.^28^ Third, it is likely that some individuals who reported five COVID-19-like symptoms had illnesses that were not COVID-19, as we were unable to collect the data on the confirmed diagnosis of COVID-19 infection (e.g., diagnosis using the PCR test). However, smell and taste disorders, one of the outcomes we used, are known to be highly specific (90% specificity) for the COVID-19 diagnosis,^15,26^ suggesting these symptoms would be good proxies of the incidence of COVID-19. Moreover, symptom-based measures would supplement the PCR test-based surveillance to understand a population-level picture of COVID-19 infection,^16,17^ because PCR testing will underestimate the true number of infections because not everyone with symptoms indicative of COVID-19 is tested. Fourth, our findings may be affected by the possibility that individuals who presented with COVID-19-like symptoms might recall and report using the subsidy program for domestic travel (as the cause of their symptoms) compared with those individuals without such symptoms (recall bias). Finally, because our study sample was collected through the web-based survey, our findings may not be generalizable to the population with limited access/literacy to the internet. Nevertheless, we used weighted analysis to minimize the difference in demographics, SES, and health-related characteristics between respondents of the current internet survey and the nationally representative survey, and thus would approximate our estimates to national estimates.

## CONCLUSION

Using a large-scale, concurrent, nationwide internet survey in Japan, we found that the participants in the government subsidies for domestic travel in Japan had higher incidence rates of COVID-19-like symptoms compared to the non-participants. Our findings suggest the implementation of the subsidy program for domestic travel might have contributed to the increased cases of the COVID-19 infection. In the midst of economic recession due to the COVID-19 pandemic, economic stimulus policies should incentivize individuals with low-risk of the COVID-19 infection to engage in economic activities while encouraging high-risk individuals to stay at home.

## Data Availability

The datasets generated during and/or analysed during the current study are available from the corresponding author on reasonable request.

## Author Contributions

Dr. Miyawaki had full access to the data in the study and takes responsibility for the accuracy and integrity of the data and its analyses.

Study concept and design: Miyawaki and Tsugawa.

Acquisition, analysis, or interpretation of data: All authors.

Drafting of the manuscript: Miyawaki and Tsugawa.

Critical revision of the manuscript for important intellectual content: All authors.

Statistical analysis: Miyawaki.

Administrative, technical, or material support: All authors.

Study supervision: Tsugawa.

