## Supplementary for "Association between Participation in Government Subsidy Program for Domestic Travel and Symptoms Indicative of COVID-19 Infection"

**Supplementary Methods 1.** Inverse Probability Weighting

**Supplementary Table 1.** Association between Participation in the Subsidy Program for Domestic Travel and Incidence of COVID-19-Like Symptoms, using the restricted sample after excluding those who were living in Tokyo

**Supplementary Table 2.** Association between Participation in the Subsidy Program for Domestic Travel and Incidence of COVID-19-Like Symptoms, Using the Unweighted Logistic Regression Models

**Supplementary Table 3.** Association between Participation in the Subsidy Program for Domestic Travel and Incidence of COVID-19-Like Symptoms, Stratified by Age

**Supplementary Table 4.** Association between Participation in the Subsidy Program for Domestic Travel and Incidence of COVID-19-Like Symptoms, Stratified by the Presence of Comorbidities

**Supplementary Table 5.** Association between Participation in the Subsidy Program for Domestic Travel and Incidence of COVID-19-Like Symptoms, for Men and Women

**Supplementary Methods 1.** Inverse Probability Weighting

Internet surveys have several advantages over traditional surveys, but the potential disadvantage is that they may not be representative of the population of interest because the subpopulations with internet access may be specific. Previous studies have used inverse probability weighting (IPW) (derived from propensity scores calculated by a logistic regression model using basic demographic and socio-economic factors such as education and length of home-ownership) obtained from an Internet-accessible convenience sample and the nationally-representative sample. It has been suggested that the parameter estimates calculated using IPW are similar or at least less different than the population-based estimates (Schonlau et al., *Sociological Methods & Research*, 2009).

In the current study, we used a population-based sample representative of the Japanese population from the 2016 Comprehensive Survey of Living Conditions (CSLC) in order to correct for sample selectivity in the internet survey. The CSLC has been conducted by the Japanese Ministry of Health, Labour and Welfare (MHLW) and collects information on health-related factors, such as self-rated health and smoking behavior, every three years (https://www.mhlw.go.jp/toukei/list/20-21.html). Out of inhabited census tracts (sampling unit for the national census in 2010), 5410 were randomly sampled across Japan in 2016 for the collection of data from all household members within each census tract. Data were available for 224,208 households (response rate; 77.5%) in 2016. Data from the 2016 CSLC were used because 2019 (latest) CSLC was not yet available at the time of analysis. Data were used with permission from MHLW. CSLC has been used in several studies (Shibuya et al., *BMJ*, 2002; Fu et al., *Journal of Health Economics*, 2017; Miyawaki et al., *BMC Geriatrics*, 2020)

We pooled and combined the data from the two surveys (the current internet survey and CSLC) and ran a multivariable logistic regression model to estimate the probability of "being an Internet survey respondent," or propensity score. Propensity scores were calculated for each group stratified by gender and age (15-19, 20-29, ..., 70-79) (gender x age stratification = 14 strata). We used variables available in both surveys (the current internet survey and CSLC) as covariates for the models. For men and women aged 20-79 years, we included socio-economic status (residence area, marital status, education, and home-ownership) and health-related characteristics (self-rated health and smoking status) in the model. For men and women aged 15-19 years, we included socio-economic status (residence area, education, and home-ownership) and self-rated health in the model, because they were too young to have a different distribution of marital status, and the CSLC did not ask teenagers their smoking status. A standardized weight was used to keep the total number of respondents included constant.

**Supplementary Table 1. Association between Participation in the Subsidy Program for Domestic Travel and Incidence of COVID-19-Like Symptoms, Using the Restricted Sample after Excluding Those Who Were Living in Tokyo**

| Subsidy Program Participation | Weighted sample, No. | Weighted incidence, n (%) | Adjusted rate, %  (95%CI) | Adjusted OR  (95%CI) | Adjusted P value |
| --- | --- | --- | --- | --- | --- |
| **High Fever** | | | | | |
| Participants | 2,959 | 308 (10.4) | 4.8  (4.4, 5.3) | 1.81  (1.34, 2.45) | <0.001 |
| Non-participants | 19,604 | 584 (3.0) | 3.9  (3.8, 4.0) | Reference |  |
| **Sore Throat** | | | | | |
| Participants | 2,959 | 622 (21.0) | 17.3  (13.2, 21.4) | 1.76  (1.19, 2.61) | 0.01 |
| Non-participants | 19,604 | 2,100 (10.7) | 11.1  (10.5, 11.8) | Reference |  |
| **Cough** | | | | | |
| Participants | 2,959 | 564 (19.1) | 16.3  (12.3, 20.3) | 1.61  (1.09, 2.39) | 0.04 |
| Non-participants | 19,604 | 2,107 (10.7) | 11.1  (10.5, 11.7) | Reference |  |
| **Headache** | | | | | |
| Participants | 2,959 | 941 (31.8) | 29.6  (27.6, 31.7) | 1.25  (1.09, 1.43) | 0.004 |
| Non-participants | 19,604 | 5003 (25.5) | 25.8  (25.5, 26.1) | Reference |  |
| **Smell and Taste Disorder** | | | | | |
| Participants | 2,959 | 157 (5.3) | 2.7  (2.1, 3.3) | 1.95  (1.11, 3.33) | 0.02 |
| Non-participants | 19,604 | 267 (1.4) | 1.9  (1.7, 2.0) | Reference |  |

SES: socio-economic status. OR: odds ratio. CI: confidence interval. We analyzed 22,563 respondents after excluding 2,919 respondents living in Tokyo. See the legend of Table 3 for more details.

**Supplementary Table 2. Association between Participation in the Subsidy Program for Domestic Travel and Incidence of COVID-19-Like Symptoms, Using the Unweighted Logistic Regression Models**

| Subsidy Program Participation | Unweighted sample, No. | Unweighted incidence, n (%) | Adjusted rate, %  (95%CI) | Adjusted OR  (95%CI) | Adjusted P value |
| --- | --- | --- | --- | --- | --- |
| **High Fever** | | | | | |
| Participants | 3,306 | 111 (3.4) | 2.4  (2.0, 2.8) | 1.54  (1.20, 1.97) | 0.001 |
| Non-participants | 22,176 | 331 (1.5) | 1.6  (1.5, 1.7) | Reference |  |
| **Sore Throat** | | | | | |
| Participants | 3,306 | 462 (14.0) | 12.8  (11.8, 13.8) | 1.23  (1.10, 1.38) | <0.001 |
| Non-participants | 22,176 | 2,338 (10.5) | 10.7  (10.5, 10.9) | Reference |  |
| **Cough** | | | | | |
| Participants | 3,306 | 455 (13.8) | 13.4  (12.4, 14.4) | 1.23  (1.10, 1.36) | <0.001 |
| Non-participants | 22,176 | 2,489 (11.2) | 11.3  (11.1, 11.4) | Reference |  |
| Participants | 3,306 | 988 (29.9) | 27.5  (26.5, 28.5) | 1.14  (1.07, 1.22) | <0.001 |
| Non-participants | 22,176 | 5,509 (24.8) | 25.2  (25.0, 25.3) | Reference |  |
| **Smell and Taste Disorder** | | | | | |
| Participants | 3,306 | 63 (1.9) | 1.4  (1.1, 1.7) | 1.54  (1.15, 2.07) | 0.004 |
| Non-participants | 22,176 | 180 (0.8) | 0.9  (0.9, 1.0) | Reference |  |

SES: socio-economic status. OR: odds ratio. CI: confidence interval. We showed the results of the analyses using unweighted logistic regression models. See the legend of Table 3 for more details.

**Supplementary Table 3. Association between Participation in the Subsidy Program for Domestic Travel and Incidence of COVID-19-Like Symptoms, Stratified by Age**

|  | Age < 65 yrs (n=19,174) | | | Age ≥ 65 yrs (n=6,308) | | |
| --- | --- | --- | --- | --- | --- | --- |
|  | Adjusted rate, % (95%CI) | Adjusted OR (95%CI) | Adjusted P value | Adjusted rate, %  (95%CI) | Adjusted OR  (95%CI) | Adjusted P value |
| **High Fever** | | | | | | |
| Participants | 6.1 (5.5, 6.8) | 2.01 (1.47, 2.74) | <0.001 | 0.8 (-0.5, 2.1) | 1.04 (0.16, 6.68) | 0.96 |
| Non-participants | 4.7 (4.6, 4.8) | Reference |  | 0.8 (0.7, 0.9) | Reference |  |
| **Sore Throat** | | | | | | |
| Participants | 23.4 (18.4, 28.4) | 2.29 (1.53, 3.43) | <0.001 | 8.3 (3.1, 13.4) | 1.24 (0.48, 3.20) | 0.65 |
| Non-participants | 12.6 (11.8, 13.5) | Reference |  | 7.1 (6.7, 7.5) | Reference |  |
| **Cough** | | | | | | |
| Participants | 21.6 (16.1, 27.1) | 2.18 (1.38, 3.44) | 0.003 | 8.1 (4.5, 11.6) | 0.80 (0.44, 1.45) | 0.46 |
| Non-participants | 11.8 (10.9, 12.7) | Reference |  | 9.7 (9.3, 9.9) | Reference |  |
| **Headache** |  |  |  |  |  |  |
| Participants | 35.6 (33.2, 38.0) | 1.27 (1.10, 1.46) | 0.001 | 10.5 (7.0, 13.9) | 1.23 (0.75, 2.03) | 0.42 |
| Non-participants | 30.9 (30.5, 31.3) | Reference |  | 9.0 (8.7, 9.3) |  |  |
| **Smell and Taste Disorder** | | | | | | |
| Participants | 3.4 (2.7, 4.1) | 2.00 (1.15, 3.48) | 0.01 | 0.3 (0, 0.6) | 0.52 (0.20, 1.38) | 0.19 |
| Non-participants | 2.4 (2.2, 2.6) | Reference |  | 0.6 (0.6, 0.6) | Reference |  |

We stratified the respondents by age (15-64 years and 65-79 years) and separately repeated the analyses using the same models as the main analyses. For Holm-adjusted P values, we multiplied the *i*-th smallest unadjusted P values by (5 – *i* + 1) times if the unadjusted P value < 0.05, and simply showed the unadjusted P values if ≥ 0.05. P for interaction (Wald test, not adjusted for multiple testing) between the subsidy program participation and age group were 0.28 for high fever, 0.09 for sore throat, 0.006 for cough, 0.18 for headache, and 0.02 for smell and taste disorder, respectively. See the legend of Table 3 for more details.

**Supplementary Table 4. Association between Participation in the Subsidy Program for Domestic Travel and Incidence of COVID-19-Like Symptoms, Stratified by the Presence of Comorbidities**

|  | Individuals without comorbidities (n=12,749) | | | Individuals with comorbidities (n=12,733) | | |
| --- | --- | --- | --- | --- | --- | --- |
|  | Adjusted rate, %  (95%CI) | Adjusted OR  (95%CI) | Adjusted P value | Adjusted rate, %  (95%CI) | Adjusted OR  (95%CI) | Adjusted P value |
| **High Fever** | | | | | | |
| Participants | 2.7 (1.7, 3.7) | 2.67 (1.58, 4.51) | <0.001 | 7.2 (6.6, 7.7) | 1.30 (0.89, 1.90) | 0.18 |
| Non-participants | 1.0 (0.9, .12) | Reference |  | 6.7 (6.6, 6.9) | Reference |  |
| **Sore Throat** | | | | | | |
| Participants | 11.6 (9.8, 13.5) | 1.36 (1.10, 1.68) | 0.02 | 25.9 (18.8 33.0) | 2.55 (1.43, 4.53) | 0.005 |
| Non-participants | 9.0 (8.7, 9.2) | Reference |  | 13.8 (12.4, 15.1) | Reference |  |
| **Cough** | | | | | | |
| Participants | 10.6 (8.5, 12.6) | 1.31 (1.01, 1.69) | 0.08 | 25.6 (18.1, 33.1) | 2.24 (1.26, 3.98) | 0.02 |
| Non-participants | 8.3 (8.1, 8.6) | Reference |  | 14.3 (12.9, 15.6) | Reference |  |
| **Headache** |  |  |  |  |  |  |
| Participants | 31.8 (28.8, 34.8) | 1.40 (1.17, 1.67) | <0.001 | 26.2 (23.5, 28.9) | 1.05 (0.85, 1.29) | 0.67 |
| Non-participants | 25.6 (25.3, 26.0) | Reference |  | 25.5 (25.0, 26.0) | Reference |  |
| **Smell and Taste Disorder** | | | | | | |
| Participants | 1.5 (0.6, 2.3) | 1.87 (0.84, 4.13) | 0.13 | 4.4 (2.7, 3.1) | 2.47 (1.30, 4.69) | 0.02 |
| Non-participants | 0.8 (0.7, 1.0) | Reference |  | 2.9 (3.6, 5.2) | Reference |  |

We stratified the respondents by the presence of comorbidities and separately repeated the analyses using the same model as the main analyses. For Holm-adjusted P values, we multiplied the *i*-th smallest unadjusted P values by (5 – *i* + 1) times if the unadjusted P value < 0.05, and simply showed the unadjusted P values if ≥ 0.05. P for interaction (Wald test, not adjusted for multiple testing) between the subsidy program participation and age group were 0.06 for high fever, 0.03 for sore throat, 0.09 for cough, 0.08 for headache, and 0.67 for smell and taste disorder, respectively. See the legend of Table 3 for more details.

|  | Men (n=12,673) | | | Women (n=12,809) | | |
| --- | --- | --- | --- | --- | --- | --- |
|  | Adjusted rate, %  (95%CI) | Adjusted OR  (95%CI) | Adjusted P value | Adjusted rate, %  (95%CI) | Adjusted OR  (95%CI) | Adjusted P value |
| **High Fever** | | | | | | |
| Participants | 7.6 (6.7, 8.5) | 1.82 (1.09, 3.02) | 0.06 | 2.7 (1.6, 3.7) | 2.50 (1.43, 4.35) | 0.005 |
| Non-participants | 6.5 (6.2, 6.7) | Reference |  | 1.2 (1.1, 1.3) | Reference |  |
| **Sore Throat** | | | | | | |
| Participants | 24.7 (18.1, 31.3) | 3.54 (2.00, 6.28) | <0.001 | 13.5 (11.1, 16.0) | 1.10 (0.84, 1.43) | 0.49 |
| Non-participants | 9.9 (8.8, 11.1) | Reference |  | 12.6 (12.2, 12.9) | Reference |  |
| **Cough** | | | | | | |
| Participants | 25.4 (18.1, 32.7) | 2.76 (1.56, 4.91) | 0.002 | 11.4 (9.8, 13.0) | 1.10 (0.90, 1.33) | 0.35 |
| Non-participants | 11.9 (10.87 13.2) | Reference |  | 10.6 (10.4, 10.8) | Reference |  |
| **Headache** |  |  |  |  |  |  |
| Participants | 21.9 (18.9, 24.9) | 1.24 (0.97, 1.59) | 0.09 | 36.8 (33.6, 39.9) | 1.28 (1.07, 1.54) | 0.03 |
| Non-participants | 18.9 (18.5, 19.4) | Reference |  | 32.0 (31.6, 32.4) | Reference |  |
| **Smell and Taste Disorder** | | | | | | |
| Participants | 3.9 (3.2, 4.6) | 1.68 (0.94, 2.99) | 0.08 | 1.7 (0.7, 2.6) | 1.99 (0.90, 4.40) | 0.09 |
| Non-participants | 3.2 (3.0, 3.4) | Reference |  | 0.9 (0.8, 1.0) | Reference |  |

**Supplementary Table 5. Association between Participation in the Subsidy Program for Domestic Travel and Incidence of COVID-19-Like Symptoms, for Men and Women**

We stratified the respondents by sex and separately repeated the analyses using the same model as the main analyses. For Holm-adjusted P values, we multiplied the *i*-th smallest unadjusted P values by (5 – *i* + 1) times if the unadjusted P value < 0.05, and simply showed the unadjusted P values if ≥ 0.05. P for interaction (Wald test, not adjusted for multiple testing) between the subsidy program participation and age group were 0.78 for high fever, 0.001 for sore throat, 0.01 for cough, 0.69 for headache, and 0.35 for smell and taste disorder, respectively. See the legend of Table 3 for more details.
